# Time trends in infectious and chronic disease consultations in Dakar, Senegal: Impact of Covid-19 Sanitary Measures

**DOI:** 10.1101/2020.11.26.20239129

**Authors:** Massamba Diop, Bamba Gaye, Stéphanie Khoury, Anouk Asselin, Samuel Kingue, Roland N’Guetta, Ibrahima Bara Diop, Camille Lassale, Crystal Cene, Xavier Jouven

## Abstract

**Background:** The impact of COVID-19 sanitary measures on the time trends in infectious and chronic disease consultations in Sub-Saharan Africa remains unknown.

**Methods:** We conducted a cohort study on all emergency medical consultations over a five-year period, January 2016 to July 2020, from SOS Medecins in Dakar, Senegal. The consultation records provided basic demographic information such as age, ethnicity (Senegalese v. Caucasian), and sex as well as the principal diagnosis using an ICD-10 classification (‘infectious, ‘chronic’, and ‘other’). Firstly, we investigated how the pattern in emergency consultation differed from March to July 2020 compared to previous years. Secondly, we examined any potential racial/ethnic disparities in COVID-19 consultation.

**Findings:** Data on emergency medical consultations were obtained from 53,583 patients of all ethnic origins. The mean age of patients was 37.0 ± 25.2 and 30.3 ± 21.7 in 2016-2019 and 45.5 ± 24.7 and 39·5 ± 23.3 in 2020 for Senegalese and Caucasians. The type of consultations between the months of January and July were similar from 2016 and 2019; however, in 2020, there was a drop among the numbers of infectious disease consultations, particularly from April to May 2020 when sanitary measures for COVID-19 were applied (average of 366.5 and 358.25 in 2016-1019 and 133 and 125 in 2020). The prevalence of chronic conditions remained steady during the same period (average of 381 and 394.75 in 2016-2019 and 373 and 367 in 2020). In a multivariate analysis after adjusting to age and sex, infectious disease consultations were significantly more likely to occur in 2016-2019 compared to 2020 (OR for 2016= 2.39, 2017= 2.74, 2018= 2.39, 2019= 2.01). Furthermore, the trend in the number of infectious and chronic consultations were similar among Senegalese and Caucasian groups, indicating no disparities among those seeking treatment.

**Interpretation:** During the implementation of COVID-19 sanitary measures, infectious disease rates dropped as chronic disease rates stayed stagnant in Dakar. Furthermore, no racial/ethnic disparities were observed among the infectious and chronic consultations.

**Key Points:** *Question:* How has the application of COVID-19 sanitary measures affected emergency medical consultations from March to July 2020 compared to previous years?

*Finding:* The rates of infectious diseases decreased as rates of chronic diseases stayed stagnant with the application of sanitary measures. Among the infectious and chronic disease consultations, no racial/ethnic disparities were observed.

*Meaning:* Understanding the effects of the sanitary measures against COVID-19 in Sub-Saharan Africa has helped emphasize the possibility of limiting the spread of other infectious diseases in this part of the world where they are still highly prevalent and the efficiency of controlling the spread of the virus while avoiding racial/ethnic disparities.

## Introduction

The severe acute respiratory syndrome coronavirus 2 (SARS-CoV-2) and the disease it causes, coronavirus disease 2019 (COVID-19), is an emerging health threat.^1^ Europe and the United States of America appear to not have used optimal strategies until the virus had spread to thousands of people. Specifically, the EU had waited to implement a COVID-19 response until 6 weeks after the first case was reported.^1^ In the US, there was a large gap in a federal COVID-19 response, allowing the virus to spread to all 50 states in a matter of 2 months since the first case was declared.^2^

With minimal resources available, Sub-Saharan Africa was poorly armed against COVID-19 and was projected to have worse outcomes due to the pandemic compared to developed countries. However, the reverse outcome was observed. Africa has been affected only moderately by COVID-19 until the end of June 2020, after which a new outbreak was observed in most African countries. As of 17 September 2020, the virus had spread to all 54 countries in Africa and has reached a total of about 1,373,926 confirmed cases and 33,255 estimated deaths.^3^ Some hypotheses have been proposed to explain the observed difference, such as lower age demographics, higher temperatures, and efficient lockdown implementation.^4^ Although sanitary measures in Sub-Saharan Africa seemed to help with controlling the number of COVID-19 cases, limited research have been done on the effectiveness of such measures and how preventative measures affect the dynamic of the pandemic and the morbidity and mortality of other prevalent diseases in Sub-Saharan Africa. Furthermore, not only has the lack of early intervention led to an unprecedent number of cases and mortality but also has shown large disparities among different racial/ethnic groups in the western countries.^5^ In several states in the US, COVID-19 mortality was highest among Latinos (187 per 100 000) and African Americans (184 per 100 000) ^6^, and in the UK, people from Black or mixed background showed higher COVID-19 infection and mortality (PHE report).

Using a prospective evaluation, we looked at all emergency medical consultations through a major emergency service provider in Dakar, Senegal (“SOS Medecins”), from January 2016 to July 2020. Firstly, we investigated the effectiveness of the sanitary measures in Africa. We hypothesized that if sanitary measures were effective in controlling COVID-19 then they should also reduce the transmission of other infectious diseases. Secondly, we assessed whether or not there were disparities between African (Senegalese) and Caucasians subjects observed in Africa like the in the US and Europe.

## Methods

### Data

We extracted data on all emergency medical consultations over a 4.5-year period (from 1^st^ January 2016 to 31 July 2020) using records of SOS Medecins, the largest provider of such services in Senegal. This service was established in 1997, and it maintains rigorous standards, with an ISO9001 international quality certification. SOS Medecins Dakar is operational at all times (24 hours a day, 7 days a week) and comprises 8 emergency physicians, 11 anaesthesiologists and 4 cardiologists. It offers mobile emergency consultations and ambulance transport with a physician or paramedic. A standardised protocol is used to document each consultation; details and a provisional diagnosis are entered into the SOS Medecins database by the attending doctor. Three days later, a final diagnosis is ascertained using additional data (phone-call to discharged patients and review of medical record). The SOS Medecins database is a robust source for information on patients who seek emergency care for diverse medical conditions in Dakar.

### Statistical analyses

For this study, data for all consecutive medical consultations handled by SOS Medecins in Dakar were obtained by a dedicated team of physician researchers. Comprehensive demographic information, including age, ethnicity (Senegalese vs Caucasian), sex and instigator of the consultation (family or patient) were collected. For our analyses, the final principal diagnosis with an ICD-10 classification were then assigned to three broad groups: 1) ‘infectious’ (Respiratory infections, gastroenteritis/diarrhea, ORL/stomato/infectious, and other infectious diseases); 2) ‘Chronic diseases (Cardiovascular disease, Rheumatological, neurological and psychiatric conditions, non-communicable non-infective pulmonary and gastrointestinal disorders); 3) Others (mainly trauma and accidents/injuries).

Data was obtained from patients of all ethnic origins. We first examined and plotted the unadjusted rates of consultations for the three disease groups between January and July from 2016 to 2020. We also analyzed separately for women and men as well as across age groups (< 20; from 20 to 45 and > 45 years of age). We then assessed and compared the unadjusted rates of the two main disease categories « infectious diseases » and « chronic diseases » from April to July (the corresponding period of the COVID-19 outbreak) for each year between 2016 and 2020.

Multivariate Logistic regression analysis was used to model the prevalence of infectious disease consultations (compared to chronic disease) across periods. Odds ratios (OR) and 95% confidence interval (CI) were calculated as estimates of the relative risk of examination for infectious disease associated with each period. All statistical analyses were performed using R version 3.6.1.

## Results

The study flowchart is detailed in **Supplemental Figure 1**. A total of 53,583 medical consultations were undertaken by SOS Medecins in Dakar over the 5-year observation period. Consultations with missing and/or incomplete data on sex of the patient (n=18), age (n=92), ethnicity (n=57) and diagnosis for the consultation (n=119) were excluded from the analyses. Complete data was obtained from a total of 53,297 consultations. **Figure 1** shows the distribution of emergency medical consultations by month from January to July over the 5-year period. Demographic data are shown in **Table 1**. The mean age of patients was 37.0 ± 25.2 and 30.3 ± 21.7 years in 2016-2019 and 45.5 ± 24.7 and 39.5 ± 23.2 years in 2020 for Senegalese and Caucasian, respectively. **Table 1** provides further details on the type of consultations received among the overall population, which are grouped under infectious and chronic, in 5-year bands and descriptive data on ICD10 diagnostic categories separately.

**Table 1.**
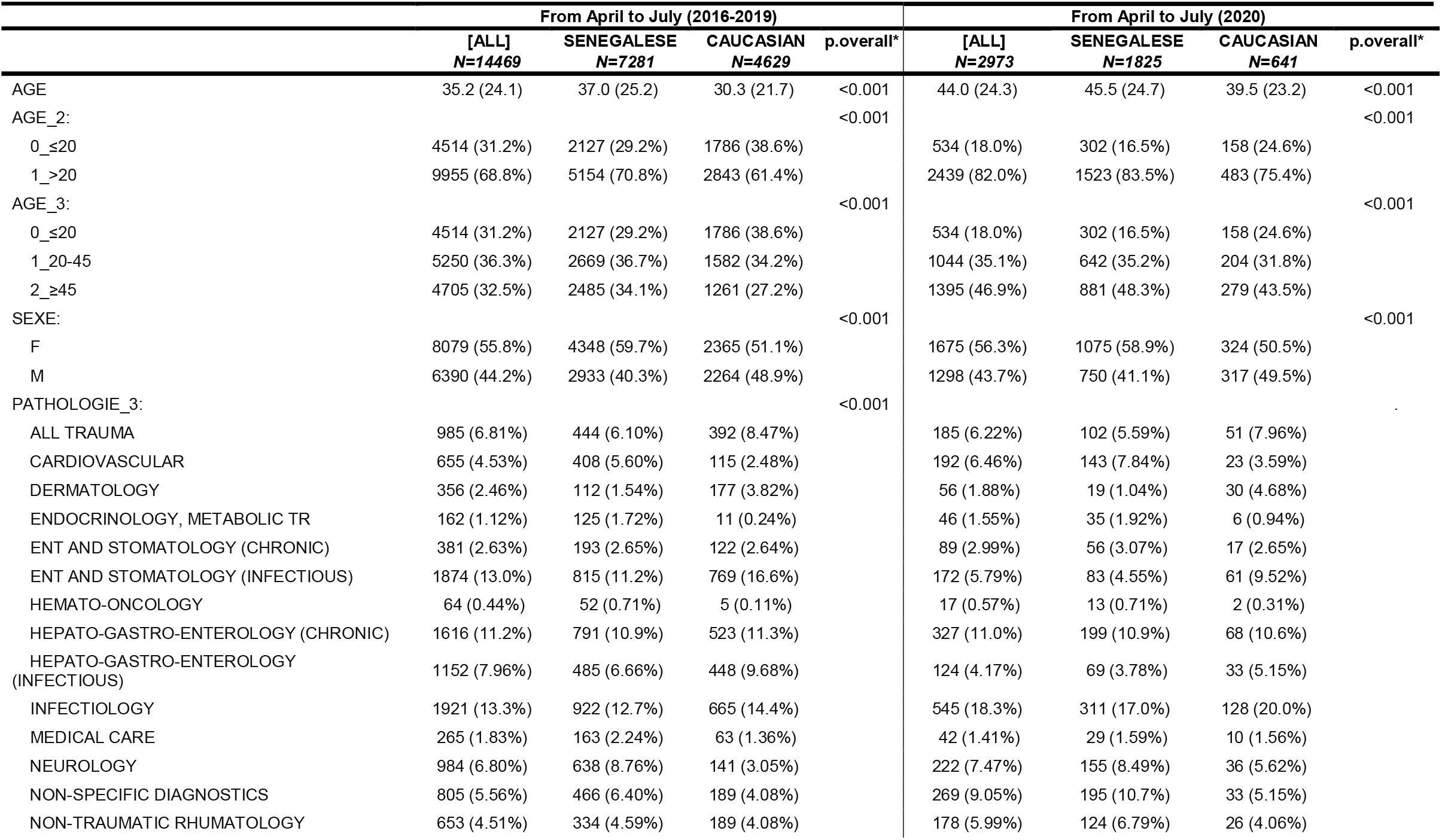
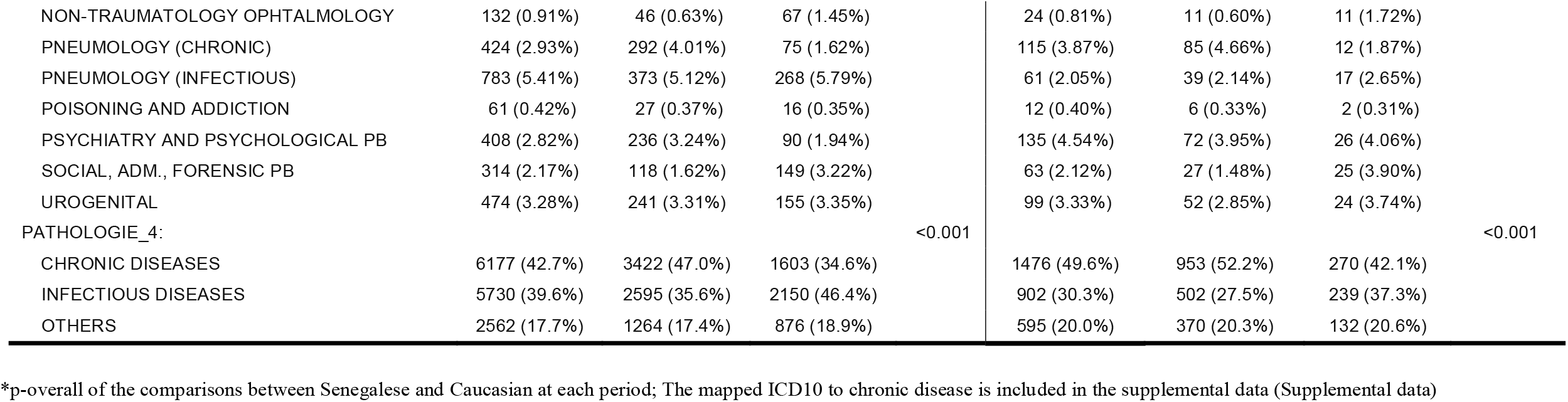
Characteristics of consultations from April to July (2020) as compared to April to July (2016-2019) in Senegalese versus Caucasian.

**Figure 1:**
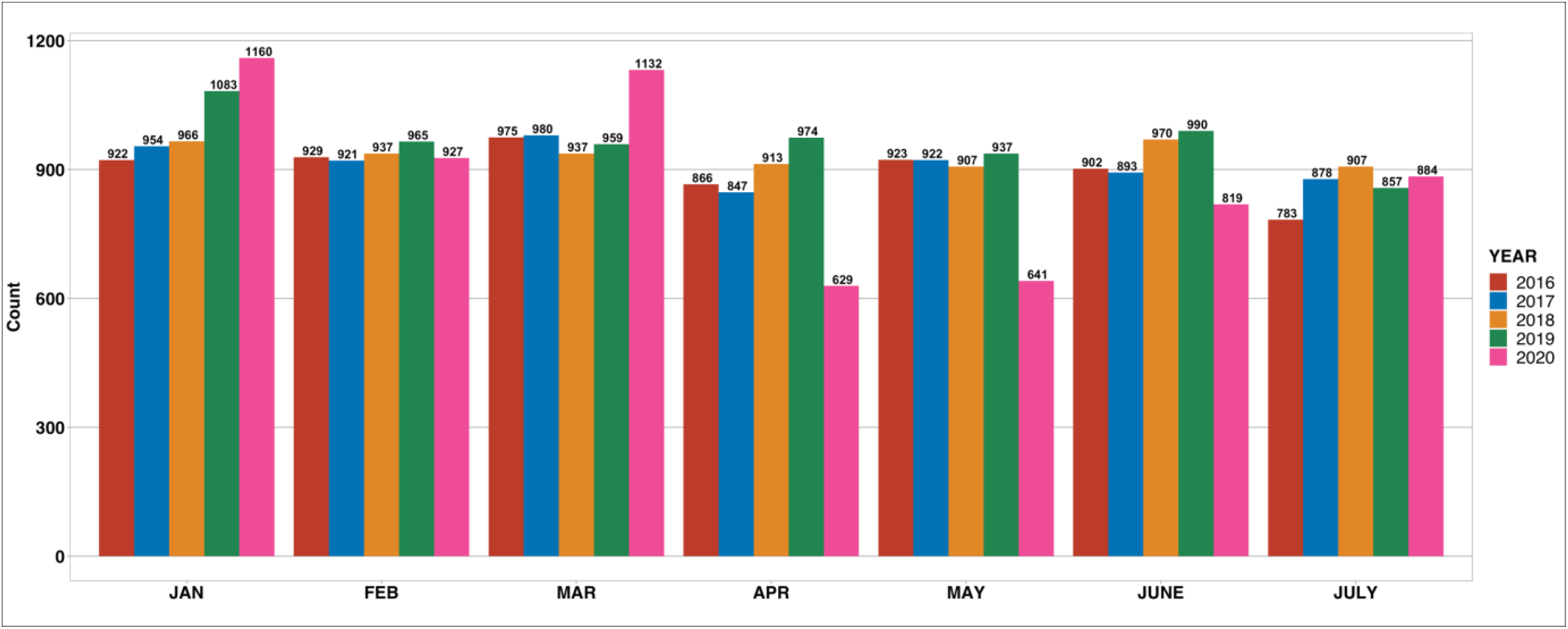
Number of emergency medical consultations by month over a 5-year period.

We first examined all consultations in the overall population grouped under infectious and chronic consultations categorized per month over 5 years. The pattern of trend in the primary cause of consultations between January and July remain similar between 2016 and 2019 (**Figure 2**, Panels A and B, respectively). In 2020, the aggregate data shows a drop in infectious diseases from April (starting-date for the application of COVID-19 prevention measures, especially quarantines measure) to May, as compared to the same period the last 4 years (**Figure 2, Panel A**) (366.5 and 358.25 in April and May 2016-2019 compared to 133 and 125 in 2020). In contrast, the corresponding prevalence of chronic conditions remain stable from over the study period (**Figure 2, Panel B**) (381 and 394.75 in April and May 2016-2019 compared to 373 and 367 in 2020). A similar and consistent pattern was observed across age groups and gender **(Supplemental Figure 2, Supplemental Figure 3)**. In a stratified analysis by subtype of infectious and chronic diseases (**Supplemental Figure 4**) the data showed a sharp decline in consultations for infections consultations in particular for Ear Nose and Throat (ENT)-Stomatology while non-communicable diseases remained stable, with the exception of psychiatry and psychological consultations that rose in April-May 2020 compared to previous years.

**Figure 2:**
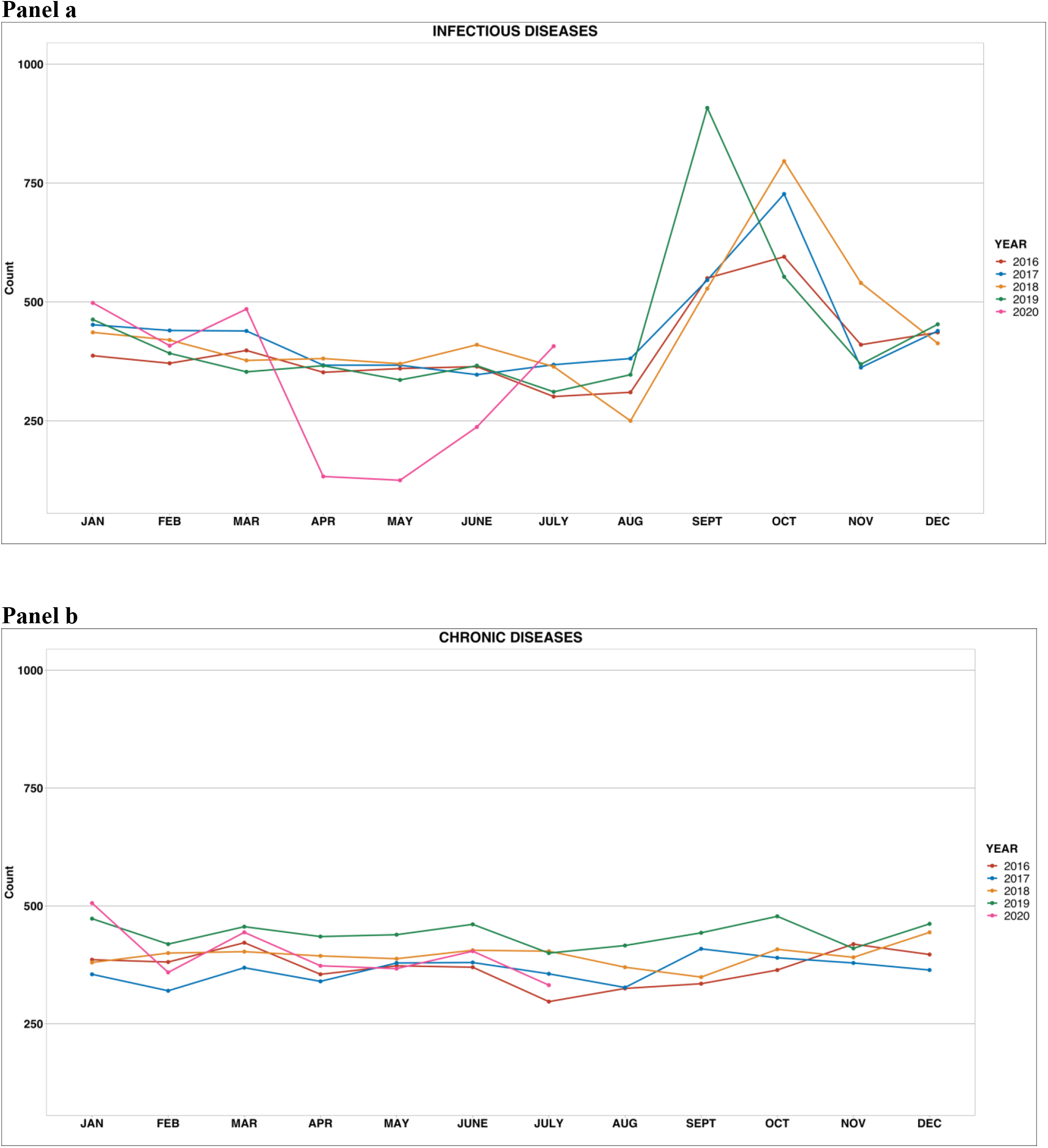
Broad diagnostic categories of emergency medical consultations by Infectious diseases (panel a) and Chronic diseases (panel b) over 5 years.

We then examined consultations between April and July in the overall Dakar population stratified by ethnicity group. The pattern of trend in the primary cause of consultations between April and July were similar across both ethnic groups between 2016 and 2020 (**Table 1, Figure 3**, Panels A and B, respectively). **Supplemental Table 1** present a comparison of the characteristics of the consultations from April-May (2020) as compared to June-July (2020) in Senegalese versus Caucasian.

**Figure 3:**
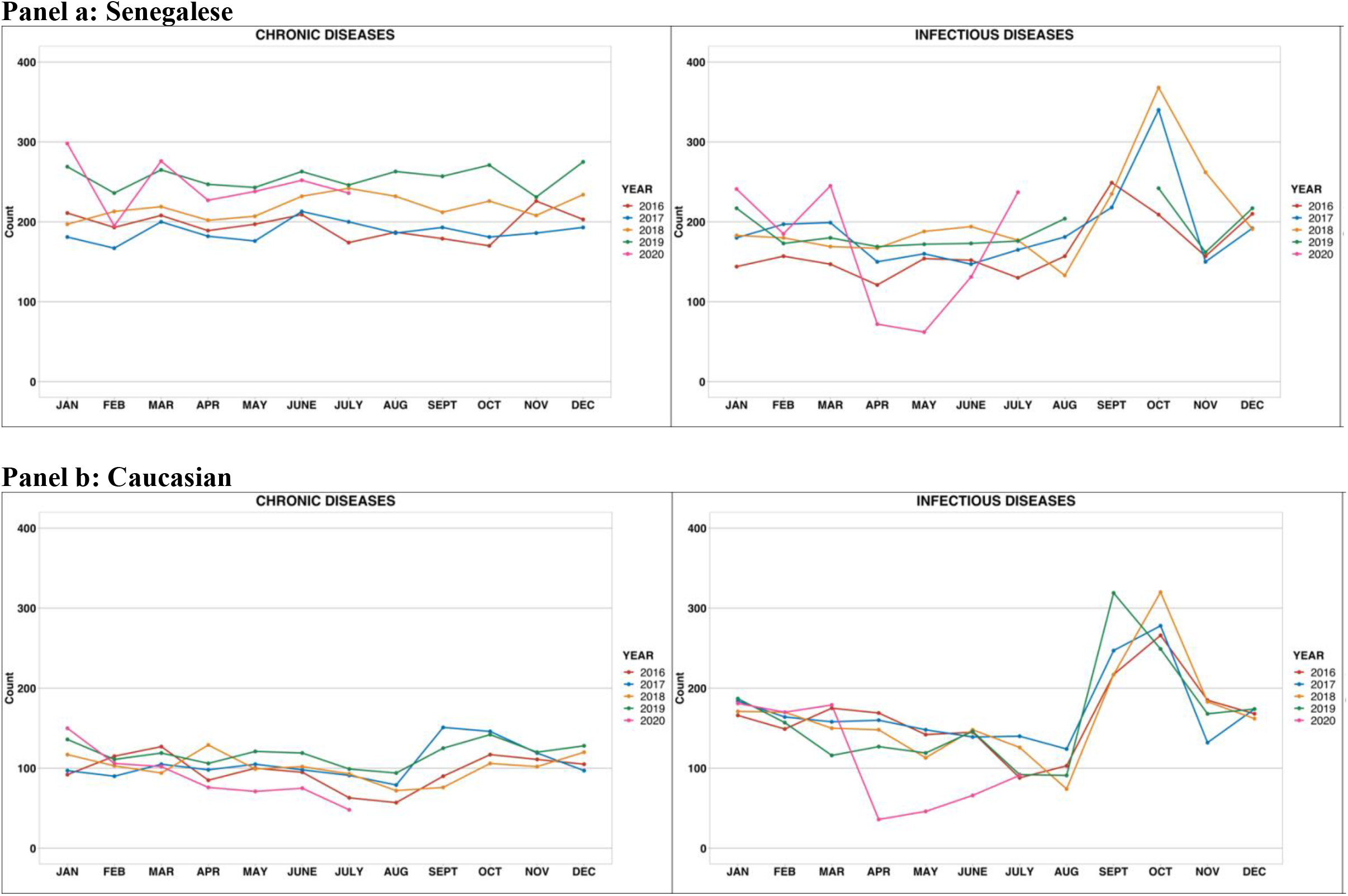
Broad diagnostic categories of emergency medical consultations by Racial/Ethnic groups: Senegalese (panel a) and Caucasian (panel b) over 5 years.

In the Multivariable logistic regression analysis after adjustment for age and sex, **Table 2** showed that consultations for infectious diseases were significantly more likely to occur in April and May 2016 to 2019 as compared to the same period in 2020 [(OR for 2016: 2.39; 95% CI: 1.96 to 2.93), (OR for 2017: 2.74; 95% CI: 2.25 to 3.36), (OR for 2018: 2.39; 95% CI: 1.96 to 2.92) and (OR for 2019= 2.01; 95% CI 1.65 to 2.45)].

**Table 2.** Multivariable Odds Ratios for excess risk of infectious diagnosis in April-May 2020 as compared to the same period in 2016, 2017, 2018 and 2019.

|  | **OR\* (95% CI)** | **p-value** |
| --- | --- | --- |
| **2016** | 2.39 (1.96-2.93) | <0.0001 |
| **2017** | 2.74 (2.25-3.36) | <0.0001 |
| **2018** | 2.39 (1.96-2.92) | <0.0001 |
| **2019** | 2.01 (1.65-2.45) | <0.0001 |
| **SEX (Men)** | 1.14 (1.02-1.28) | 0.02135 |
| **† Age = 20-45** | 0.38 (0.33-0.43) | <0.0001 |
| **Age ≥ 45** | 0.21 (0.18-0.24) | <0.0001 |

\* Logistic regression adjusted for age and sex., N= 5721; 2020 as the reference exposure category; † reference age is < 20 years old.

As presented in Table 3, and after adjustment for age and sex, there was a significant increasing rate of consultation for infectious diagnosis in June and July as compared to April and May 2020.

**Table 3.**
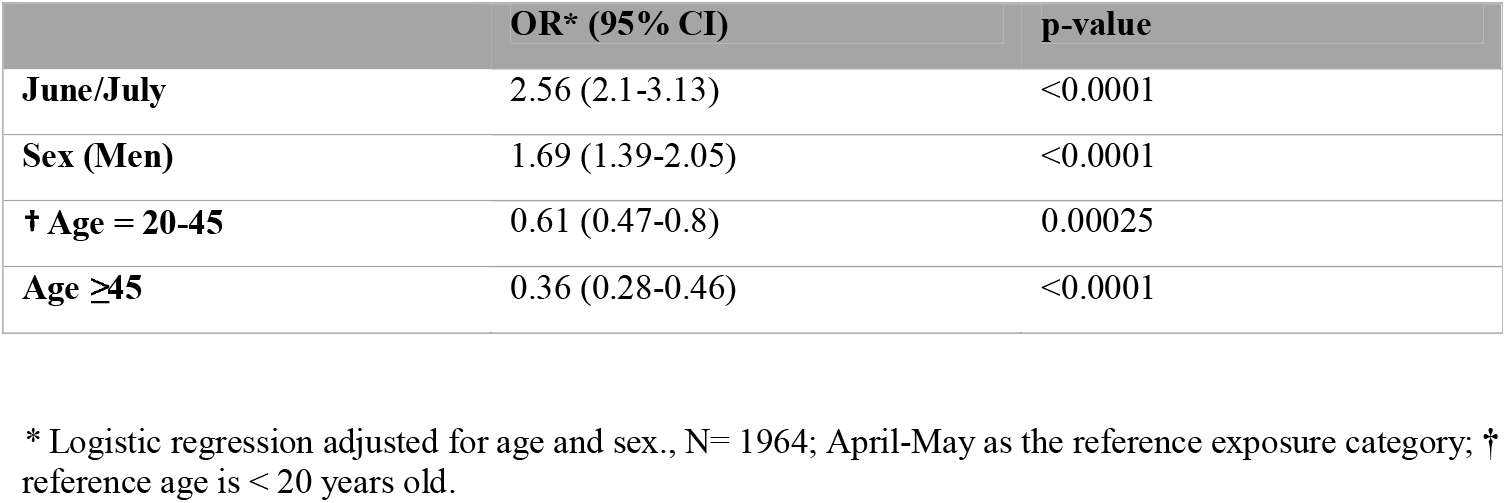
Multivariable Odds Ratios for excess risk of infectious diagnosis in June and July as compared to April and May 2020.

## Discussion

In this study, using data on consultations in a large, well-documented emergency medical service in Dakar, we reported two key findings. Firstly, we observed a decrease in consultation for infectious diseases starting from the application of COVID-19 sanitary measures (April-May 2020) as compared to the same period over the previous 5 years, while the number of consultations for chronic diseases remained stable. Secondly, in our study, we do not report any disparities across ethnic groups in terms of number of consultations for both infectious and chronic disease.

During the period covering the implementation of COVID-19 sanitary measures (April to May 2020), infectious disease consultations dropped significantly while chronic diseases remained stable. During an outbreak such as COVID-19, one would assume consultations for infectious disease rates would increase as the virus spreads throughout the country. However, as it has been previously reported, COVID-19 has not affected countries in Africa as dramatically as the westernized world.^4^ The decrease in infectious disease cases found in our study is a first step toward understanding the low spread of the COVID-19 disease in Africa. Our finding suggests that sanitary measures may have limited the spread of COVID-19 and other infectious diseases.

Each country implemented swift preventative measures that contributed to controlling the spread of the virus. Such prevention methods included border control, travel restriction, the use of face coverings/masks and social distancing (maintaining a distance of one meter from others), social media campaigns, quarantines, and emphasizes on proper sanitation and hygiene practices.^7^ A content-wide response, the Africa Taskforce on Coronavirus Preparedness and Response (AFTCOR), was also carried out.^8^ The AFTCOR was formed by the African Center for Disease Control and Prevention (Africa CDC) and the Southern Africa Center for Infectious Disease Surveillance (SACIDS).^9^ Such centralized response included surveillance in high-risk countries and provided proper laboratory testing and diagnosis; both strategies were deemed successful during the 2014 and 2018 Ebola epidemics.^9,4^

It is also important to note that cases of infectious disease began to rise again between May and July. An explanation for this sudden increase could be that sanitary measures were no longer enforced as strictly as they once were at the beginning of the outbreak. The loosened COVID-19 restrictions have led people to take less precautions and therefore, increase rates of transmission of infectious diseases, including COVID-19. Furthermore, this finding carries large public health implications as such strategies to limit the spread of the COVID-19 can be used to combat any other potential epidemics that plague Sub-Saharan Africa, in particular Ebola, tuberculosis or diarrheal diseases.

Another possible contribution to the decline in infectious disease cases is that people may have been more hesitant to go to the health facility because of the fear of catching the virus, especially if they were experiencing mild disease. Chronic diseases such as CVD and other NCDs are more severe at times and require continuous monitoring, possibly explaining why chronic disease consultations did not decrease when sanitary measures were applied in Senegal. In contrast, many consultations and appointments were postponed due to COVID-19 in Europe.^10^ It is predicted that the UK will see an estimated increase of about 18,000 deaths in a year due to delayed diagnosis and treatment of cancer patients during the pandemic.^10^ In France, 38% and 28% of patients reported canceling of medical services due to the fear of infection and disturbing physicians during a pandemic.^10^

Surprisingly, in our study, consultations for chronic diseases during the COVID-19 outbreak remain stable as compared to the previous 5 years. In fact, evidence shows that social isolation is expected to take a toll on a populations mental and physical health, especially during an outbreak such as this one. The COVID-19 sanitary measures required people to stay inside their homes, leaving many unable to work or go to school. We observed a higher rate of consultation for psychiatry during the first months of implementation of COVID-19 hygienic measures. Although research on the effects of sanitary measures on mental health during COVID-19 is limited, studies on previous epidemics have shown an increase in psychiatric symptoms^11^ as well as cardiac conditions ^12^ during and after control measures were implemented. A study on the Ebola outbreak of 2014 showed the prevalence of anxiety-depression and PTSD was 48% and 76% due to the Ebola response.^11^ Similar findings were identified during the SARS and H1N1 outbreaks.^11^

In our study, there was no disparities in morbidity or mortality during the COVID-19 period across racial/ethnic unlike the US and the UK.^13^ Blacks and other minority groups face a higher rate of COVID-19 cases and deaths in the US and the UK as well as an overall increased burden of chronic diseases during the pandemic.^6,14^ Several hypothesis including social determinants of health, preexisting conditions, genetics, structural racism, behavioral and economic aspects have been raised.^4^ In our study, the Senegalese and Caucasians are likely to have a quite similar socioeconomic status and our results are, therefore, in favor of socioeconomic status playing a major role in the differences observed between ethnic groups in the US and the UK. Moreover, Senegal has tackled COVID-19 aggressively and, so far, effectively leading to a very low number of COVID-19 cases and deaths. More than six months into the pandemic, the country had about 14,000 cases and 284 deaths making comparisons at the ethnic level challenging given the lack of statistical power.

Overall, this study represents unique data on the immediate effect of the sanitary measures during COVID-19 in Senegal and provides relevant information that will help to pursue further research in Senegal and other Sub-Saharan African countries.

An important limitation of this study is the use of a single database, SOS Medecins Dakar, to address our questions. It is unclear whether or not our observations reflect a direct association between COVID-19 prevention measures and its effect on consultations. Furthermore, despite the fact that this study is based on the largest database of emergency medical consultations in Senegal, the extent to which it is fully representative of healthcare utilization in emergency medicine in an urban setting is unclear as other emergency care services were not involved due to lack of documentation in these services. Data were derived from the capital city and it is possible that disease patterns and resource utilization in other regions, especially more rural areas, may be different. However, this study provides unique data on the potential effect of the sanitary measures against COVID-19 in Africa and emphasizes the need to limit the spread of all infectious diseases in this part of the world where they are still highly prevalent.

## Conclusion

In conclusion, African countries should keep implementing sanitary measures including face mask, social distancing, hand cleaning and remain vigilant in the upcoming weeks to months. Furthermore, the absence of disparities across ethnic groups open new gates in the investigation of the mechanism underlying the COVID-19 racial gap found in the UK and the US.

## Data Availability

The SOS Medecins emergency medical consultation data in Dakar, Senegal are available for request from INSERM.

## Acknowledgments

Author contributions: BG and XJ requested the data; BG and XJ designed the study; AA conducted the data analysis; BG, XJ and S Khoury drafted the manuscript; BG, XJ, CC, MD, ID, S Khoury, RN, S Kingue and CL interpreted the results; All authors read and critically revised the manuscript.

