## Supplemental for "Time trends in infectious and chronic disease consultations in Dakar, Senegal: Impact of Covid-19 Sanitary Measures"

**Supplemental Data**

- **e-Figure 1.** Flowchart of Sample Selection.
- **e-Figure 2.** Number of emergency medical consultations for chronic and infectious diseases by age group (panel a) and gender (panel b) over a 5-year period.
- **e-Figure 3.** Broad diagnostic categories of emergency medical consultations for chronic and infectious diseases by age group (Panel a) and gender (Panel b) over a 5-year period.
- **e-Figure 4.** Number of emergency medical consultations for Cardiovascular (Panel a), Non-Traumatic Rheumatology (Panel b), Psychiatry and Psychological (Panel c), and Ear Nose and Throat and Stomatology (Panel d) disease over a 5-year period
- **e-Table 1.** Characteristics of the consultations from April to May (2020) as compared to June to July (2020).

**e-Figure 1: Flow chart of Sample Selection**

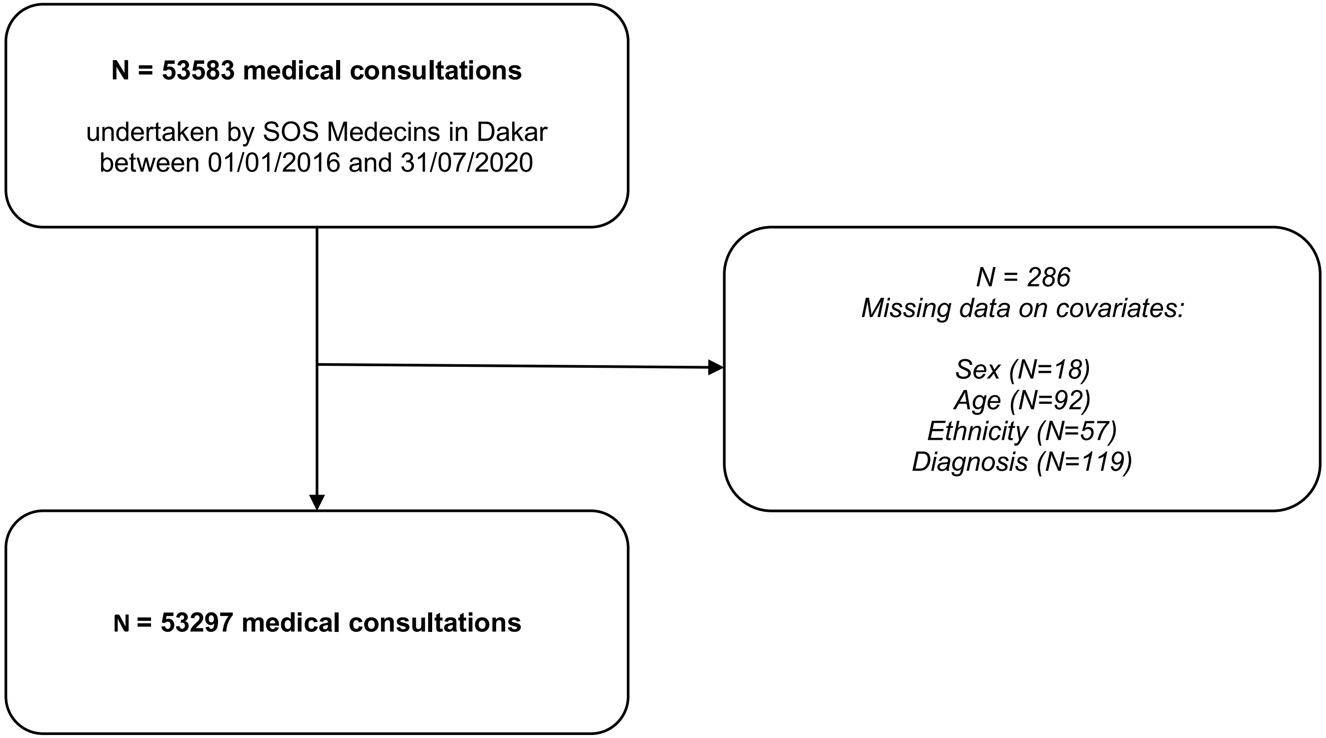

**e-Figure 2: Number of emergency medical consultations for chronic and infectious diseases by age group (panel a) and gender (panel b) over a 5-year period.**

#### Panel a

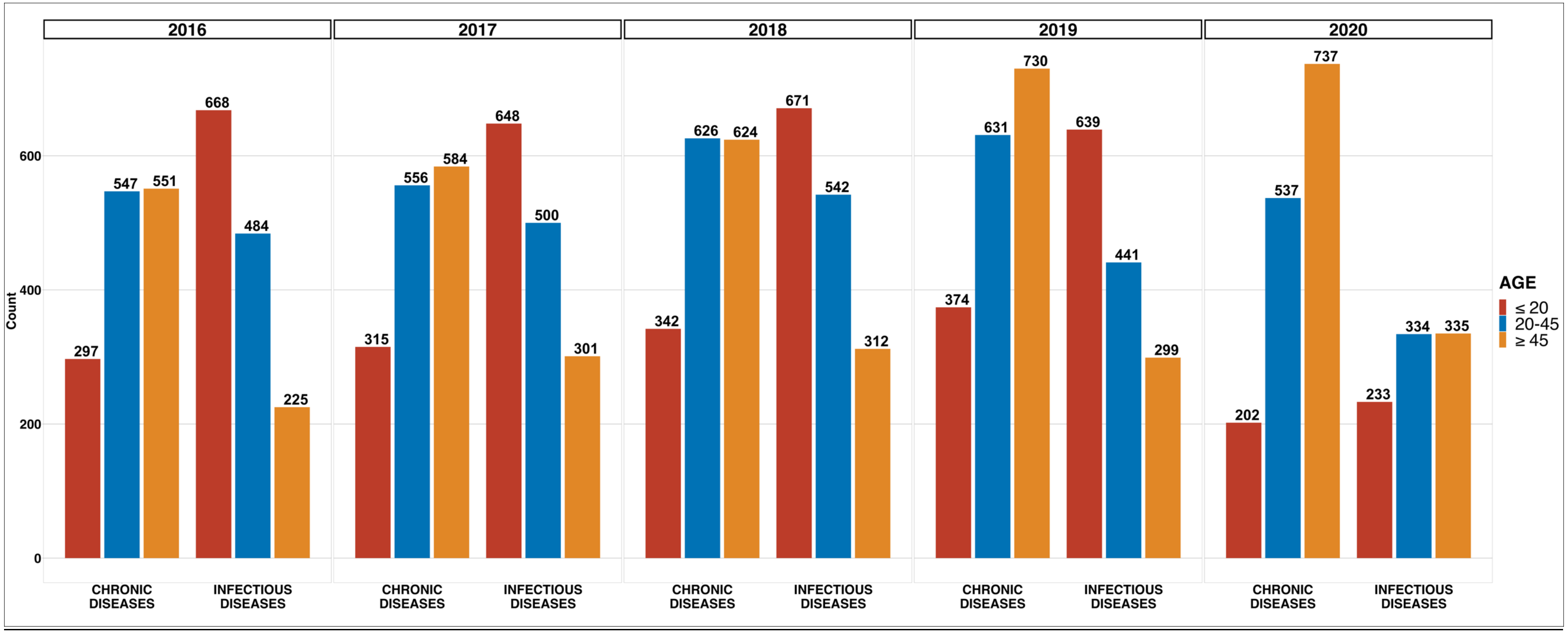

**Panel b**

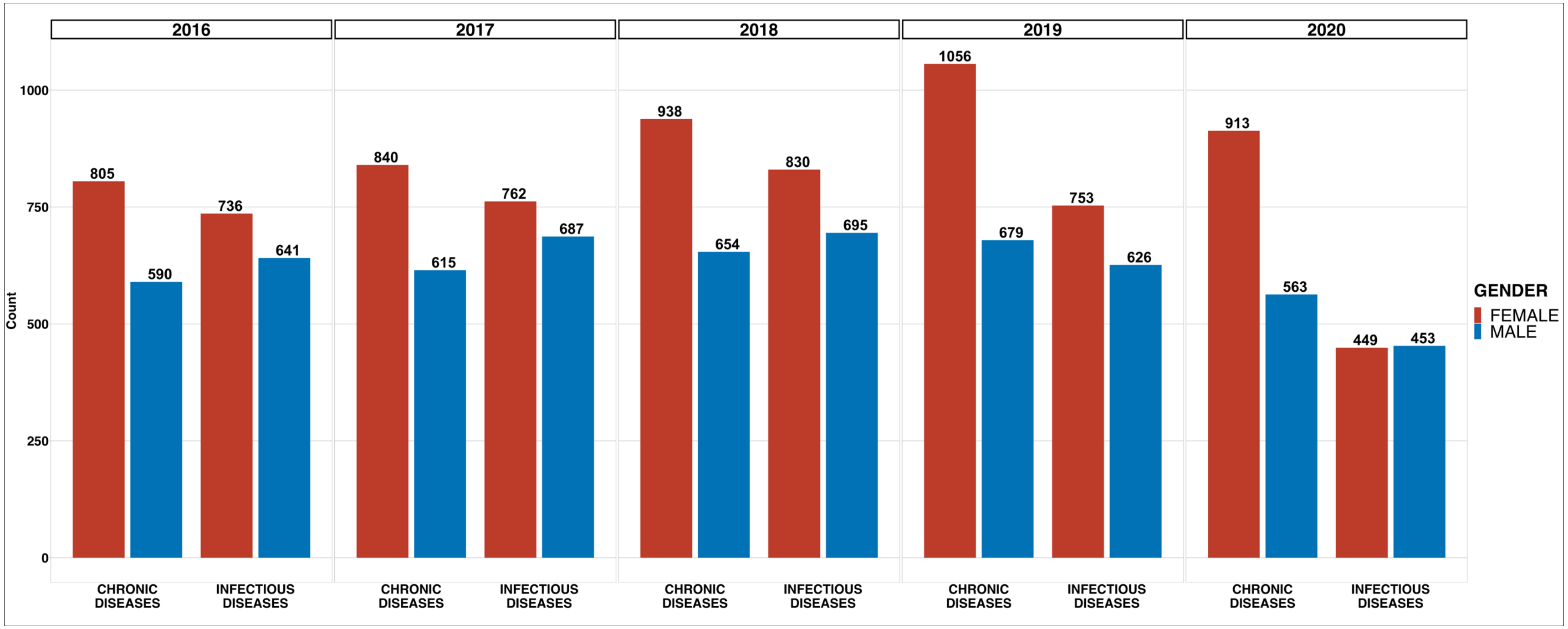

2016
2017
2018
2019
2020
Count
0
250
500
750
1000
805
590
736
641
840
615
762
687
938
654
830
695
1056
679
753
626
913
563
449
453
CHRONIC DISEASES
INFECTIOUS DISEASES
GENDER
FEMALE
MALE

**e-Figure 3: Broad diagnostic categories of emergency medical consultations for chronic and infectious diseases by age group (Panel a) and gender (Panel b) over a 5-year period.**

#### Panel a

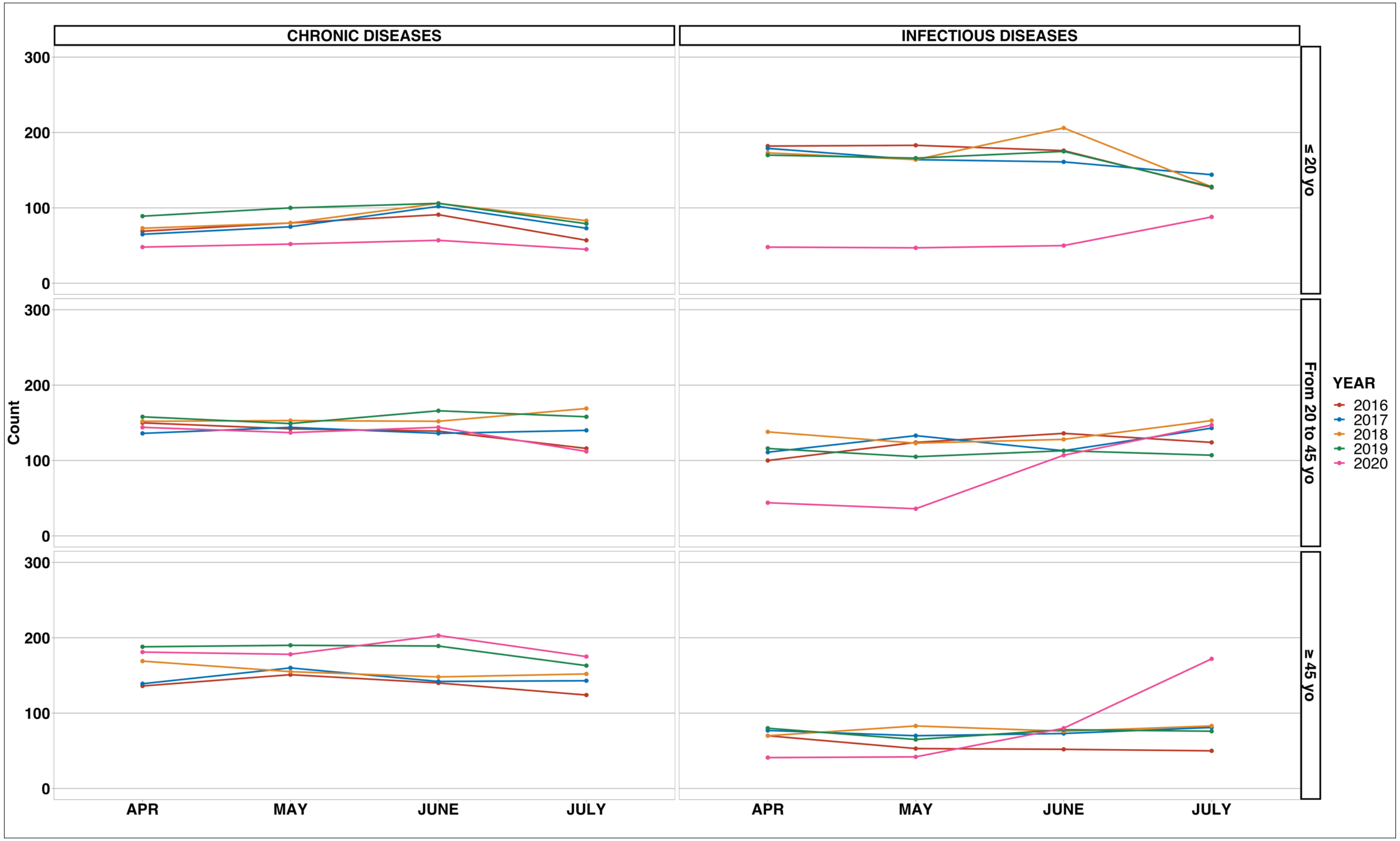

### Panel b

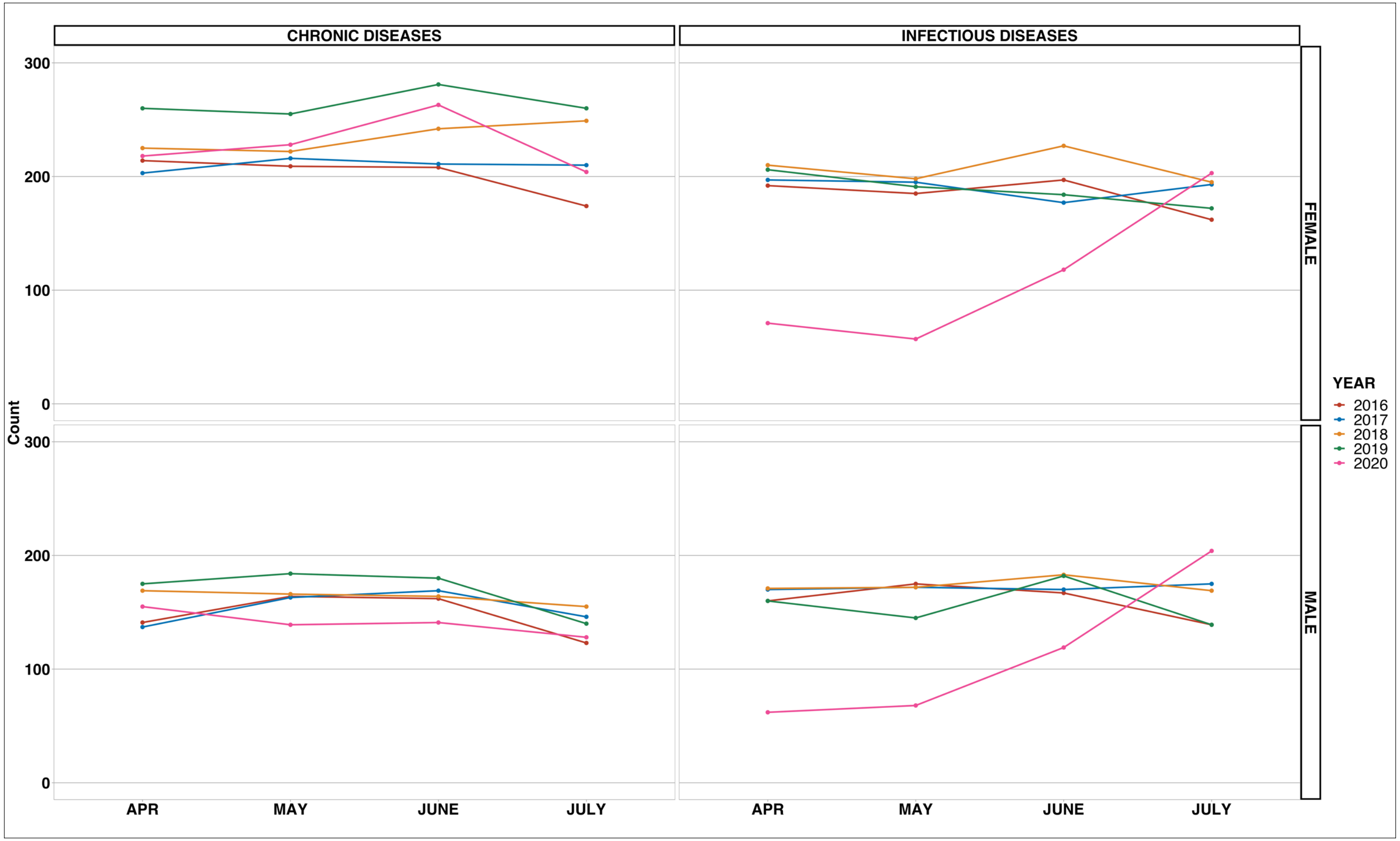
CHRONIC DISEASES
INFECTIOUS DISEASES
FEMALE
MALE
Count
300
200
100
0
APR
MAY
JUNE
JULY
YEAR
2016
2017
2018
2019
2020

**e-Figure 4: Broad diagnostic categories of emergency medical consultations by Cardiovascular (Panel a), Non-Traumatic Rheumatology (Panel b), Psychiatry and Psychological conditions (Panel c), and ENT and infectious Stomatology (Panel d) disease over a 5-year period**

#### Panel a

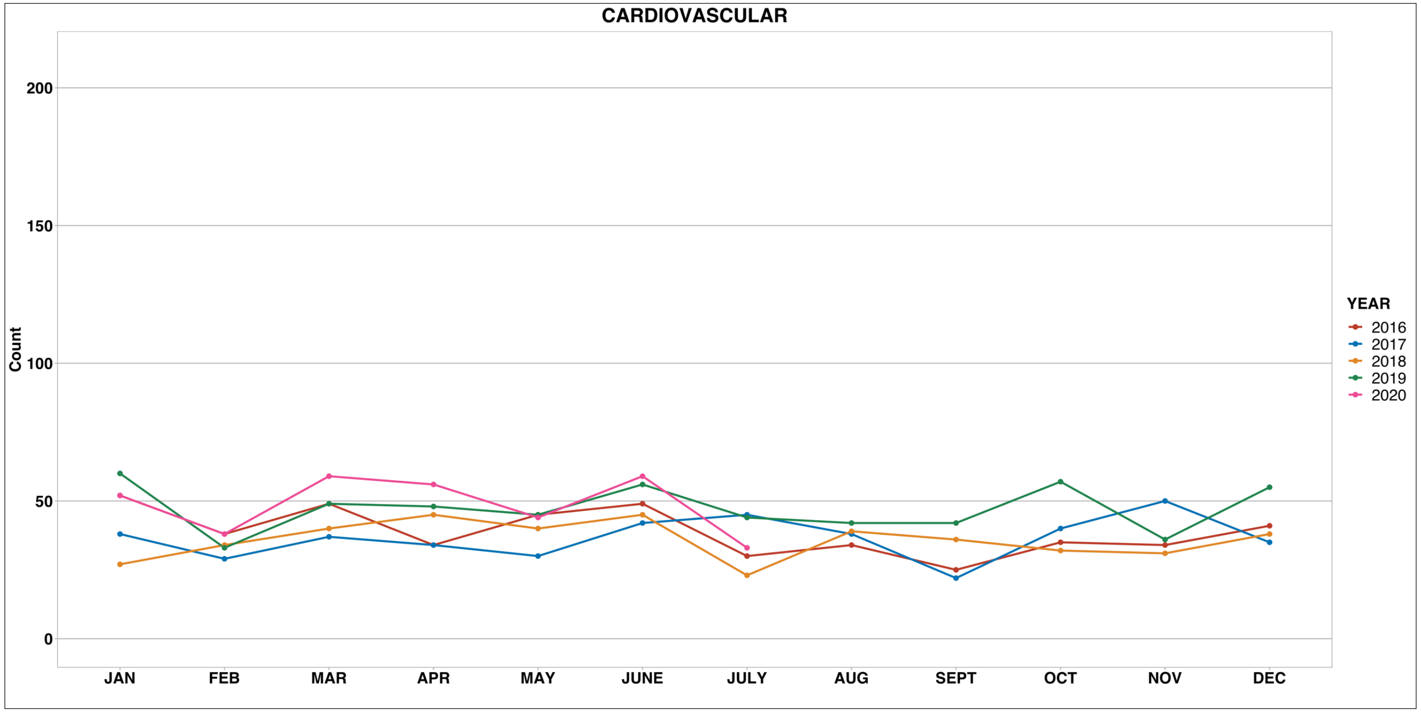

**Panel b**

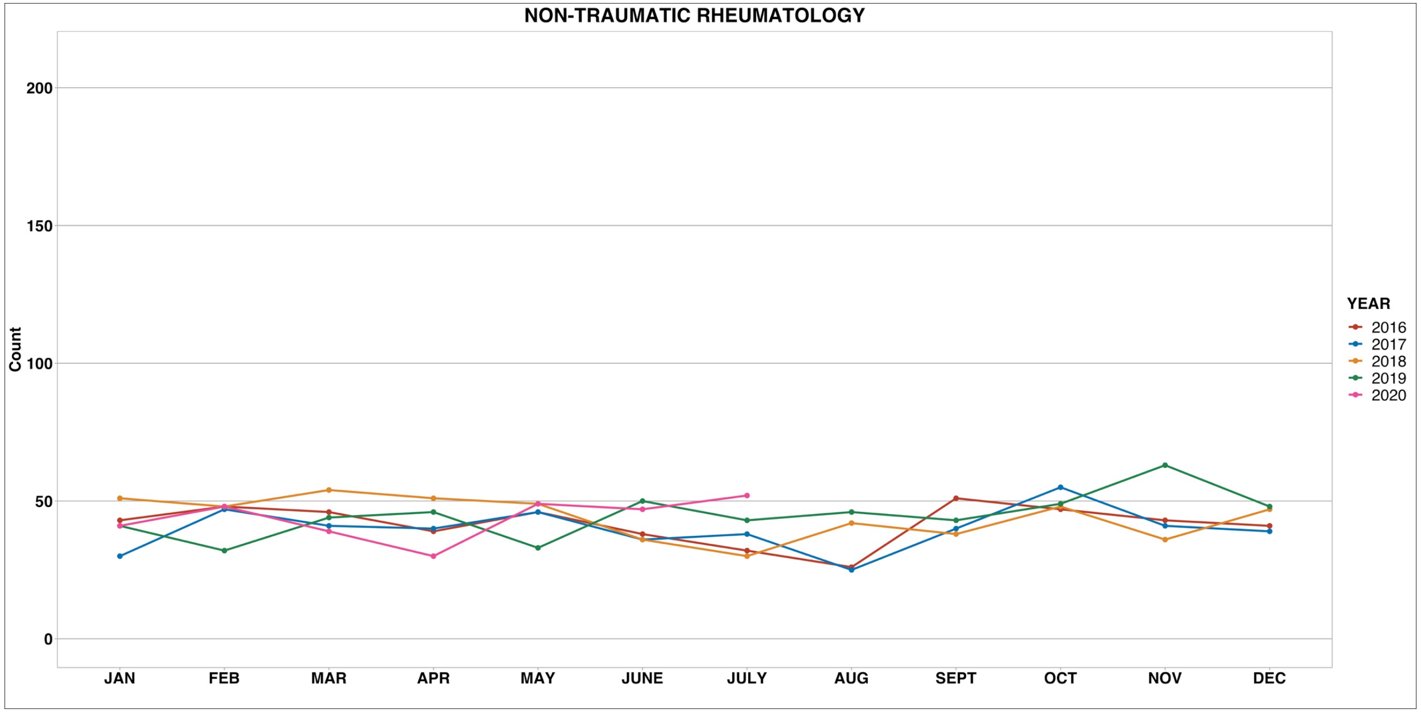

NON-TRAUMATIC RHEUMATOLOGY
Count
200
150
100
50
0
JAN
FEB
MAR
APR
MAY
JUNE
JULY
AUG
SEPT
OCT
NOV
DEC
YEAR
2016
2017
2018
2019
2020

**Panel c**

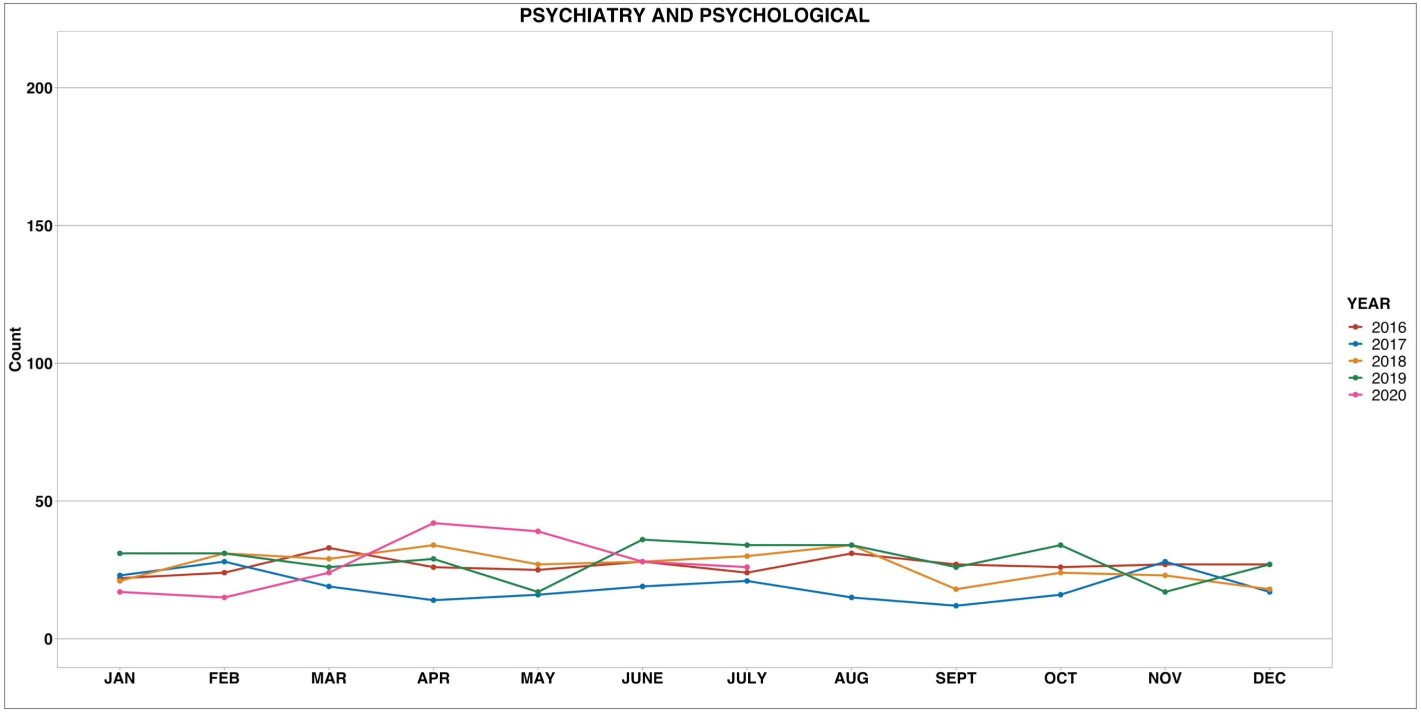

PSYCHIATRY AND PSYCHOLOGICAL
Count
200
150
100
50
0
JAN
FEB
MAR
APR
MAY
JUNE
JULY
AUG
SEPT
OCT
NOV
DEC
YEAR
2016
2017
2018
2019
2020

Panel d

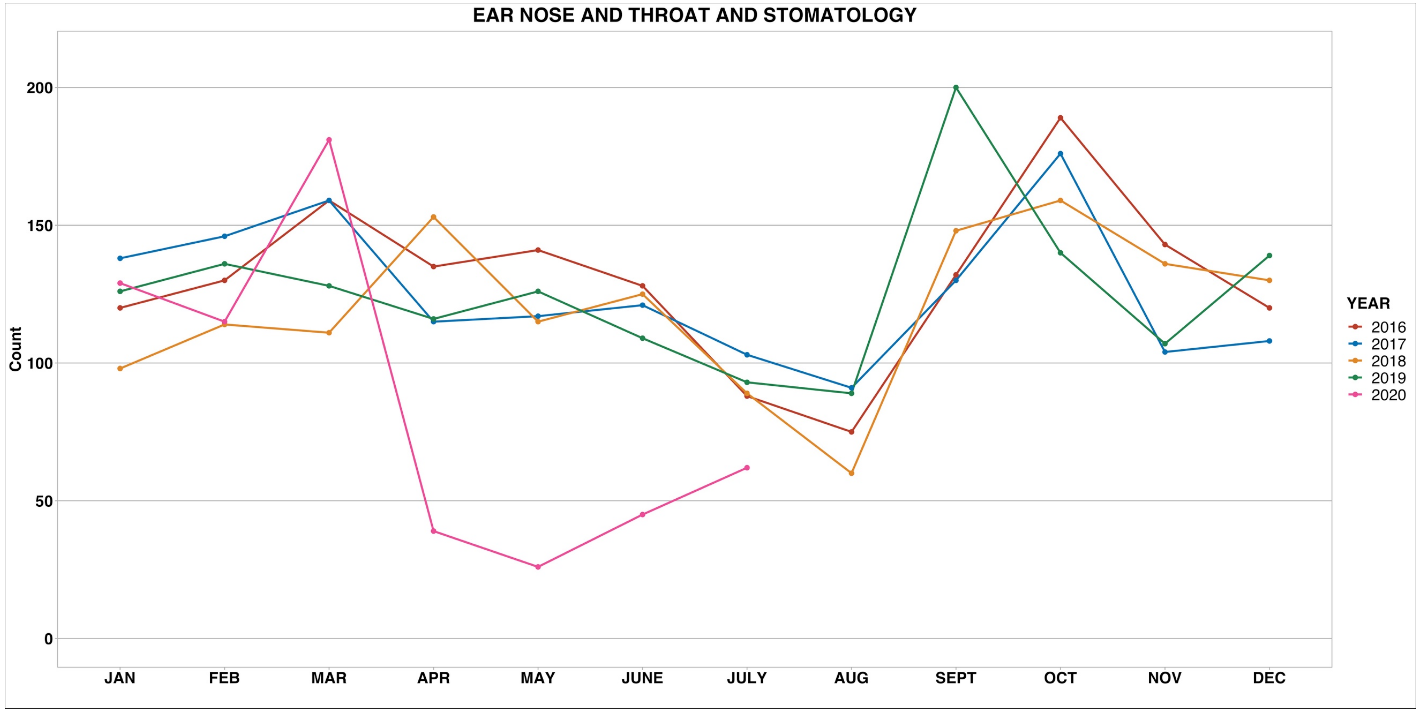

EAR NOSE AND THROAT AND STOMATOLOGY
Count
0
50
100
150
200
JAN
FEB
MAR
APR
MAY
JUNE
JULY
AUG
SEPT
OCT
NOV
DEC
YEAR
2016
2017
2018
2019
2020

**e-Table 1. Characteristics of the consultations from April to May (2020) as compared to June to July (2020).**

| | From April to May 2020 | | | | From June to July 2020 | | | |
|---|---|---|---|---|---|---|---|---|
| | [ALL] *N=1270* | SENEGALESE *N=761* | CAUCASIAN *N=305* | p.overall | [ALL] *N=1703* | SENEGALESE *N=1064* | CAUCASIAN *N=336* | p.overall |
| AGE | 42.5 (24.2) | 43.9 (24.8) | 39.1 (24.2) | 0.004 | 45.1 (24.2) | 46.6 (24.5) | 39.9 (22.4) | <0.001 |
| AGE_2: | | | | <0.001 | | | | 0.013 |
| 0_≤20 | 249 (19.6%) | 138 (18.1%) | 86 (28.2%) | | 285 (16.7%) | 164 (15.4%) | 72 (21.4%) | |
| 1_>20 | 1021 (80.4%) | 623 (81.9%) | 219 (71.8%) | | 1418 (83.3%) | 900 (84.6%) | 264 (78.6%) | |
| AGE_3: | | | | 0.001 | | | | 0.024 |
| 0_≤20 | 249 (19.6%) | 138 (18.1%) | 86 (28.2%) | | 285 (16.7%) | 164 (15.4%) | 72 (21.4%) | |
| 1_20-45 | 443 (34.9%) | 266 (35.0%) | 85 (27.9%) | | 601 (35.3%) | 376 (35.3%) | 119 (35.4%) | |
| 2_≥45 | 578 (45.5%) | 357 (46.9%) | 134 (43.9%) | | 817 (48.0%) | 524 (49.2%) | 145 (43.2%) | |
| SEXE: | | | | 0.022 | | | | 0.005 |
| F | 720 (56.7%) | 452 (59.4%) | 157 (51.5%) | | 955 (56.1%) | 623 (58.6%) | 167 (49.7%) | |
| M | 550 (43.3%) | 309 (40.6%) | 148 (48.5%) | | 748 (43.9%) | 441 (41.4%) | 169 (50.3%) | |
| PATHOLOGIE_3: | | | | . | | | | . |
| ALL TRAUMA | 100 (7.87%) | 58 (7.62%) | 25 (8.20%) | | 85 (4.99%) | 44 (4.14%) | 26 (7.74%) | |
| CARDIOVASCULAR | 100 (7.87%) | 76 (9.99%) | 15 (4.92%) | | 92 (5.40%) | 67 (6.30%) | 8 (2.38%) | |
| DERMATOLOGY | 29 (2.28%) | 8 (1.05%) | 18 (5.90%) | | 27 (1.59%) | 11 (1.03%) | 12 (3.57%) | |
| ENDOCRINOLOGY, METABOLIC TR | 18 (1.42%) | 16 (2.10%) | 1 (0.33%) | | 28 (1.64%) | 19 (1.79%) | 5 (1.49%) | |
| ENT AND STOMATOLOGY (CHRONIC) | 51 (4.02%) | 37 (4.86%) | 6 (1.97%) | | 38 (2.23%) | 19 (1.79%) | 11 (3.27%) | |
| ENT AND STOMATOLOGY (INFECTIOUS) | 65 (5.12%) | 36 (4.73%) | 19 (6.23%) | | 107 (6.28%) | 47 (4.42%) | 42 (12.5%) | |
| HEMATO-ONCOLOGY | 6 (0.47%) | 6 (0.79%) | 0 (0.00%) | | 11 (0.65%) | 7 (0.66%) | 2 (0.60%) | |
| HEPATO-GASTRO-ENTEROLOGY (CHRONIC) | 171 (13.5%) | 100 (13.1%) | 38 (12.5%) | | 156 (9.16%) | 99 (9.30%) | 30 (8.93%) | |
| HEPATO-GASTRO-ENTEROLOGY (INFECTIOUS) | 57 (4.49%) | 30 (3.94%) | 17 (5.57%) | | 67 (3.93%) | 39 (3.67%) | 16 (4.76%) | |
| INFECTIOLOGY | 117 (9.21%) | 57 (7.49%) | 39 (12.8%) | | 428 (25.1%) | 254 (23.9%) | 89 (26.5%) | |
| MEDICAL CARE | 19 (1.50%) | 11 (1.45%) | 7 (2.30%) | | 23 (1.35%) | 18 (1.69%) | 3 (0.89%) | |
| NEUROLOGY | 113 (8.90%) | 74 (9.72%) | 21 (6.89%) | | 109 (6.40%) | 81 (7.61%) | 15 (4.46%) | |
| NON-SPECIFIC DIAGNOSTICS | 109 (8.58%) | 75 (9.86%) | 21 (6.89%) | | 160 (9.40%) | 120 (11.3%) | 12 (3.57%) | |
| NON-TRAUMATIC RHUMATOLOGY | 79 (6.22%) | 54 (7.10%) | 11 (3.61%) | | 99 (5.81%) | 70 (6.58%) | 15 (4.46%) | |
| NON-TRAUMATOLOGY OPHTALMOLOGY | 14 (1.10%) | 7 (0.92%) | 6 (1.97%) | | 10 (0.59%) | 4 (0.38%) | 5 (1.49%) | |

| | | | | | | | | |
|---|---|---|---|---|---|---|---|---|
| PNEUMOLOGY (CHRONIC) | 34 (2.68%) | 28 (3.68%) | 2 (0.66%) | | 81 (4.76%) | 57 (5.36%) | 10 (2.98%) | |
| PNEUMOLOGY (INFECTIOUS) | 19 (1.50%) | 11 (1.45%) | 7 (2.30%) | | 42 (2.47%) | 28 (2.63%) | 10 (2.98%) | |
| POISONING AND ADDICTION | 6 (0.47%) | 2 (0.26%) | 2 (0.66%) | | 6 (0.35%) | 4 (0.38%) | 0 (0.00%) | |
| PSYCHIATRY AND PSYCHOLOGICAL PB | 81 (6.38%) | 41 (5.39%) | 18 (5.90%) | | 54 (3.17%) | 31 (2.91%) | 8 (2.38%) | |
| SOCIAL, ADM., FORENSIC PB | 24 (1.89%) | 9 (1.18%) | 15 (4.92%) | | 39 (2.29%) | 18 (1.69%) | 10 (2.98%) | |
| UROGENITAL | 58 (4.57%) | 25 (3.29%) | 17 (5.57%) | | 41 (2.41%) | 27 (2.54%) | 7 (2.08%) | |
| PATHOLOGIE_4: | | | | <0.001 | | | | <0.001 |
| CHRONIC DISEASES | 740 (58.3%) | 465 (61.1%) | 147 (48.2%) | | 736 (43.2%) | 488 (45.9%) | 123 (36.6%) | |
| INFECTIOUS DISEASES | 258 (20.3%) | 134 (17.6%) | 82 (26.9%) | | 644 (37.8%) | 368 (34.6%) | 157 (46.7%) | |
| OTHERS | 272 (21.4%) | 162 (21.3%) | 76 (24.9%) | | 323 (19.0%) | 208 (19.5%) | 56 (16.7%) | |
